# Normative reference values of the handgrip strength for the Portuguese workers

**DOI:** 10.1101/2020.01.21.20018333

**Authors:** Sarah Fernandes Bernardes, Ana Assunção, Carlos Fujão, Filomena Carnide

**Affiliations:** University of Lisbon, Faculty of Human Kinetics, CIPER, Sports & Health Department, Biomechanics and Functional Morphology Laboratory, Lisbon, Portugal; Volkswagen Autoeuropa – Area of Industrial Engineering and Lean Management, Palmela, Portugal; Institute of Education and Science, Universitas. Lisbon, Portugal

## Abstract

**Aim:** This study aims to identify the normative values of handgrip strength for Portuguese workers in the automotive industry.

**Methods:** About 1225 employees were invited to participate in the study. The final sample consisted of 656 employees in the assembly area. The handgrip strength was measured in kilograms (kg) using the Jamar digital dynamometer. Two measurements were performed in both hands, totaling four measurements. For the present study, the maximum value was recorded regardless of the hand.

**Results:** showed the peak mean values of handgrip strength in the group of women was 34 kg in the age group of 35-39 years, and the group of men the peak mean was 52 kg in the age group of 25-34 years. The most pronounced decline in the female group appears in the age of 30-34 years with 30 kg and the men group the decline occurs of 2kg below the peak force, in the age group between the 40-57 years. This study used a cut-off at 2 SD below by the sex-specific peak mean.

**Conclusion:** Normative values can help delineate the career path of workers because they portray risk values according to age, height, and gender. And they can also help in adjusting the morphological and strength characteristics of the worker with the task to be performed, as an example of work above head level.

**Key Messages:**

- The handgrip strength is a general indicator of muscle strength, in addition to being closely related to cardiovascular and nutritional diseases. Also, this measure is one of the keys to defining sarcopenia.
- One of the main findings of the study was found in the group of women aged 30-34 years, with a significant decline in handgrip strength compared to other age groups. Furthermore, the present study, established for the first time, normative values for the handgrip strength of Portuguese workers in the automotive industry.
- The handgrip strength decline is a crucial predictor of frailty syndrome, and sarcopenia can be checked by the occupational medicine department, individually, by the risk threshold outcomes presents in this study. Also, it is possible to design the conditions work processes associated with the predictive values of HG2 and HG5 and the implementation of the workers’ clinical surveillance system through periodic tests of handgrip strength.

## Introduction

Aging is a process of progressive decline cellular, physical, and mental capabilities (1). The phenomenon of aging and the incidence of related diseases such as sarcopenia (progressive and significant loss of muscle mass and strength) and frailty syndrome (decline of the physiological system related to age, affecting strength and endurance increasing the risk of falls, dependence and or death), may arise early and may be aggravated by exposure to work demands during the active course of professional life (2–7).

That situation occurs because the interaction with the workload, working time, and the intensity of physical exposure can reduce the muscle’s ability to generate energy, which in turn facilitates the development of these age-related syndromes (8,9).

Some measures are used to diagnose sarcopenia and frailty syndrome, such as handgrip strength (HGS), which is considered a good inter-rater reliability (4,10–12). The handgrip force is measured by the amount of static force the hand can grip a dynamometer (13) and is an indicator of overall muscle strength (14).

Factors such as age and gender are considered influencers in HGS in healthy individuals (15). About age, the tendency is that HGS decreased with increasing age and regarding gender, female HGS presents lower values (16).

Sarcopenia and frailty syndrome are rarely studied in working-age people, and research related to work disability is scarce. It is conceivable that symptoms of frailty may be useful markers for employment difficulties in late middle life. In this case, by analogy with advances in care for the elderly, there could be room to intervene and to prevent health-related job loss and premature retirement.

Strategies may be needed to identify workers at particular risk of frailty when they are older and to help them - for example, through interventions to promote their functional capacity in middle and old age. These strategies must be adjusted to the functional capacity of workers, as they get older. Therefore, this study aims to identify the normative values of handgrip strength for Portuguese workers in the automotive industry.

## Methodology

### Sample

A cross-sectional study was conducted in the final assembly area of an automobile industry. All workers were invited to participate in the study (n=1225).

For sample size calculation participants were stratified by gender and age. The women’s workers were divided into five age groups and the men’s workers in six age groups. Height measures were restricted to 146-177 cm for women and 150-190 cm for men.

In order to ensure normative values for healthy workers, the following exclusion criteria were applied: a) minimum value of 10 kg of gripping force; b) present medical restriction and / or occupational disease; c) SF-12 criteria (Short Form Health Survey) with a score lower than 5% of the physical component score of the quality of life scale (17).

The scientific committee of the Faculty of Human Kinetics from the University of Lisbon approved the study protocol. All workers were informed about the purpose and procedures of the study and gave their written informed consent.

## Methods

### SF-12 (Short Form Health Survey)

The SF-12 is composed by 12 items organized according to a Likert scale and includes physical components score (PCS) and mental components scores (MCS). The physical dimension comprises items related to physical function, physical performance, pain, and health in general, and the mental dimension covers mental health, emotional performance, social function, and vitality (17,18).

### Anthropometry

For the height measurement was obtained with an upright position, participant-centred position on the tape, in relation to the stadiometer, footwear, arms extended along the body, feet joined or slightly separated. The head was oriented according to the Frankfurt plane, parallel to the ground, regardless of the worker’s posture (19). Due to the European safety norms industry setting, to height measure were removed 3 centimetres to compensate the height of the work shoes sole.

### Handgrip Strength

For the manual grip strength test, the standard position was used for all participants (17,20,21).The grip strength was measured in kilograms (kg) using the Jamar digital dynamometer (20). Two measurements were performed in both hands. Position 2 of the Jamar dynamometer was used, because is the most appropriate position to measure the handgrip strength (20,22).The maximum value obtained with either hand is used as a summary measure of a person’s isometric strength of the hand and forearm muscles (23,24).The Jamar dynamometer is extremely used, which is validated as a gold standard, with high test-retest reproducibility (r> 0.8) and excellent reliability (r = 0.98) (12).

### Data analyses

In order to perform the final sample, the eligibility criterion will be applied, based on the SF-12 PCS score, being standardized from the z-standardized with an average value of 36 and SD 2.35, defining the criterion for 5% below the PCS average.

Descriptive statistical analysis was used to determine mean (M), standard deviation (SD) and median values (ME) of the handgrip strength, stratified by five age groups of the women and the six age groups of the men. Sex-specific profiles of handgrip strength were designed by Ordinary Least Square regression (OLS) analysis, where height, age, age squared and height squared entered in the models as determinant factors of the maximum grip strength in the women and men groups. The objective of this step was to verify the mean peak values for women and men.

After, the cut-off values were calculated in order to identify workers with weak grip strength. These values were defined as 1SD and 2SD below the mean peak value and stratified by gender. The resulting values were plotted from the age groups defined by height.

Finally, handgrip strength measurements were first standardized for age and body height. It was determined the risk threshold, subtracting 1 SD from the mean value of height-adjusted handgrip strength and age group. The standardized measures of handgrip strength values were the z-standardized residuals derived from sex-specific OLS regressions of handgrip strength values in kg on age (in years) and body height (in cm).

## Results

The sample was composed by 656 workers that accepted to participate in the study. From those, 39 workers were excluded because they don’t fulfil the eligibility criteria, namely the 5% the physical component score (PCS) based in SF-12, This criterion was chosen to restrict the test population to healthy employees only. In addition to the SF-12 standards, it also verified the list of employees who had any medical restriction from the occupational health department of the automotive industry.

So, the final sample integrated 617 workers, mainly men (∼74%), with a mean age of 33±8,58 years and mean height of 173±6,50 cm; Women presented mean age of 32±8,03 years and mean height of 160±5,95 cm.

From the OLS regression, the mean peak of the grip strength was obtained. The mean peak value of handgrip strength for women was 34 kg and was reached in the age group of 35-39 years (Fig 1) in contrast for men that showing the mean peak value of handgrip strength of about 52 kg in the age group of 25-34 years (Fig 2.)

**Figure 1.**
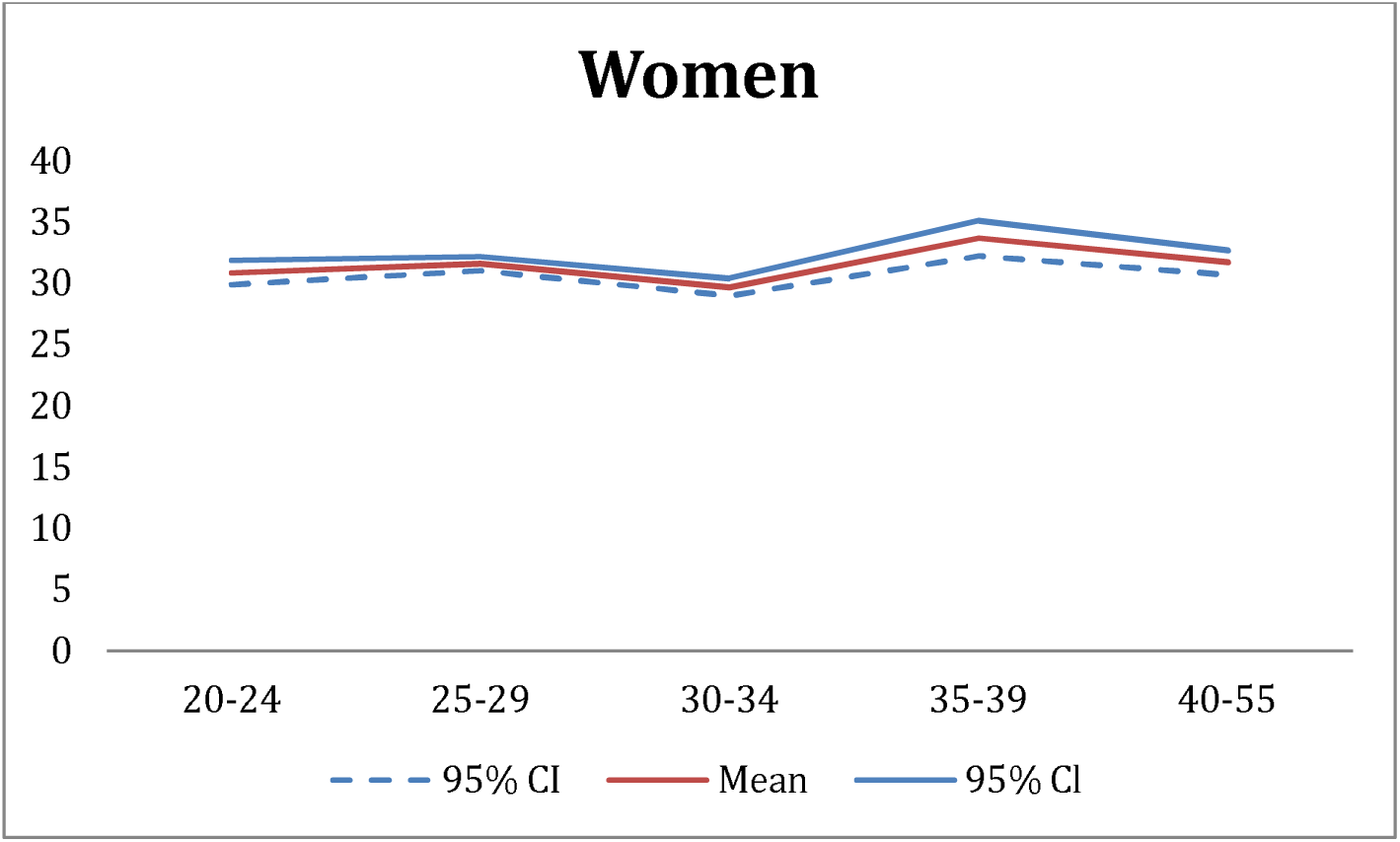
Average peak of grip strength from the OLS regression in the female group.

**Figure 2.**
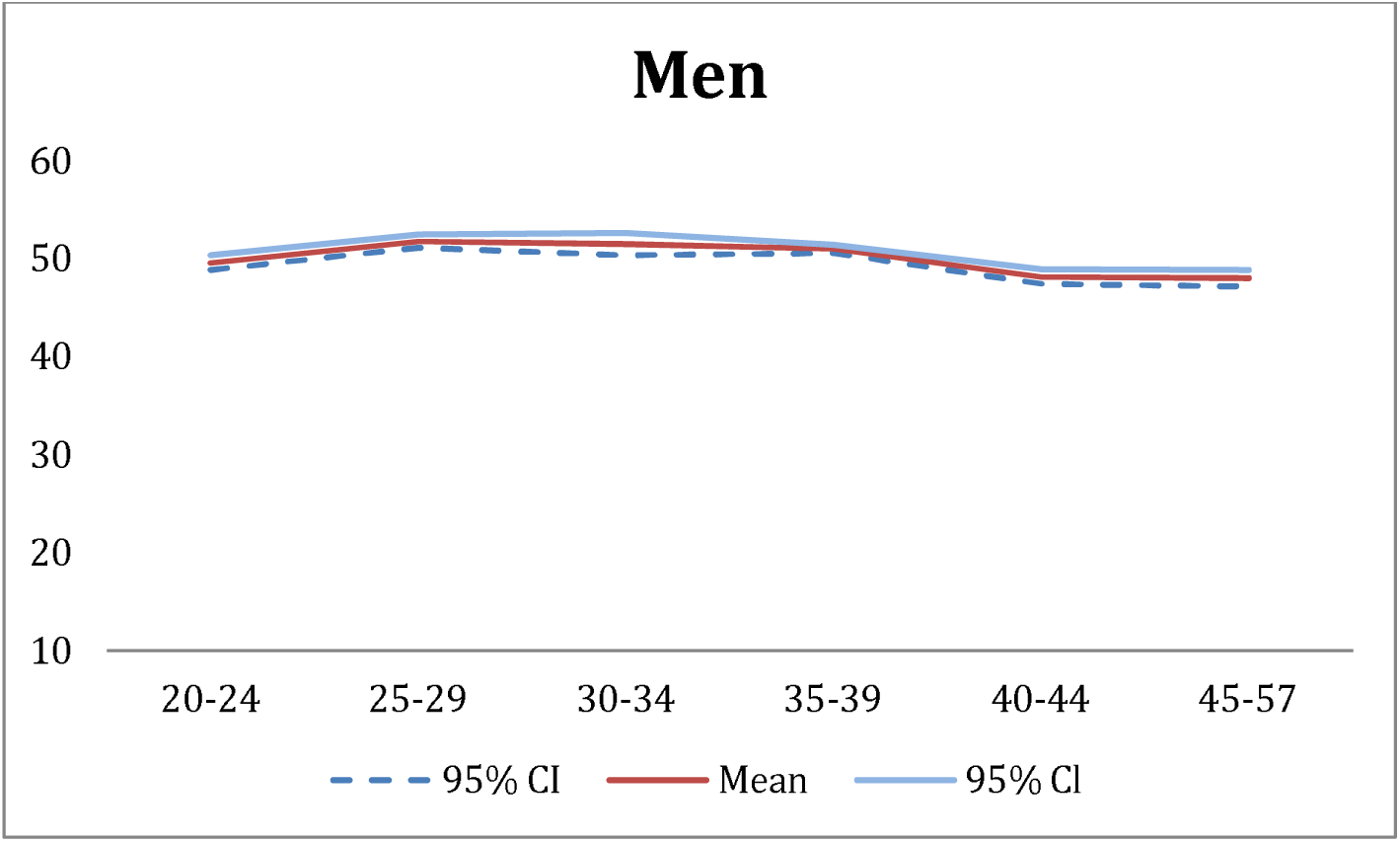
Mean peak values of handgrip strength extracted from the OLS regression in the men workers.

The most pronounced decline appears in women in the age group of 30-34 years, with 4kg (fig 1.) below the mean peak strength. For the men, this decline occurs in 2kg, in the age group between the 40-57 years (fig. 2).

Simultaneously, it was determined, the prevalence of workers in each of the weak handgrip strength, through 1 SD and 2 SD below the peak mean values also in relation to the low grip strength cut-off for men at 27 kg and for women 16 kg, based on (4). The age group of 20-24 in the women’s group had the values of 1 SD and 2 SD, respectively, 25.4 kg and 20.1 kg. In the age group of 30-34 years in the group of women, they had the lowest values of deviations, and in the 2 SD (15.1Kg), they were below the cut-off (see fig 3a).

**Figure 3a.**
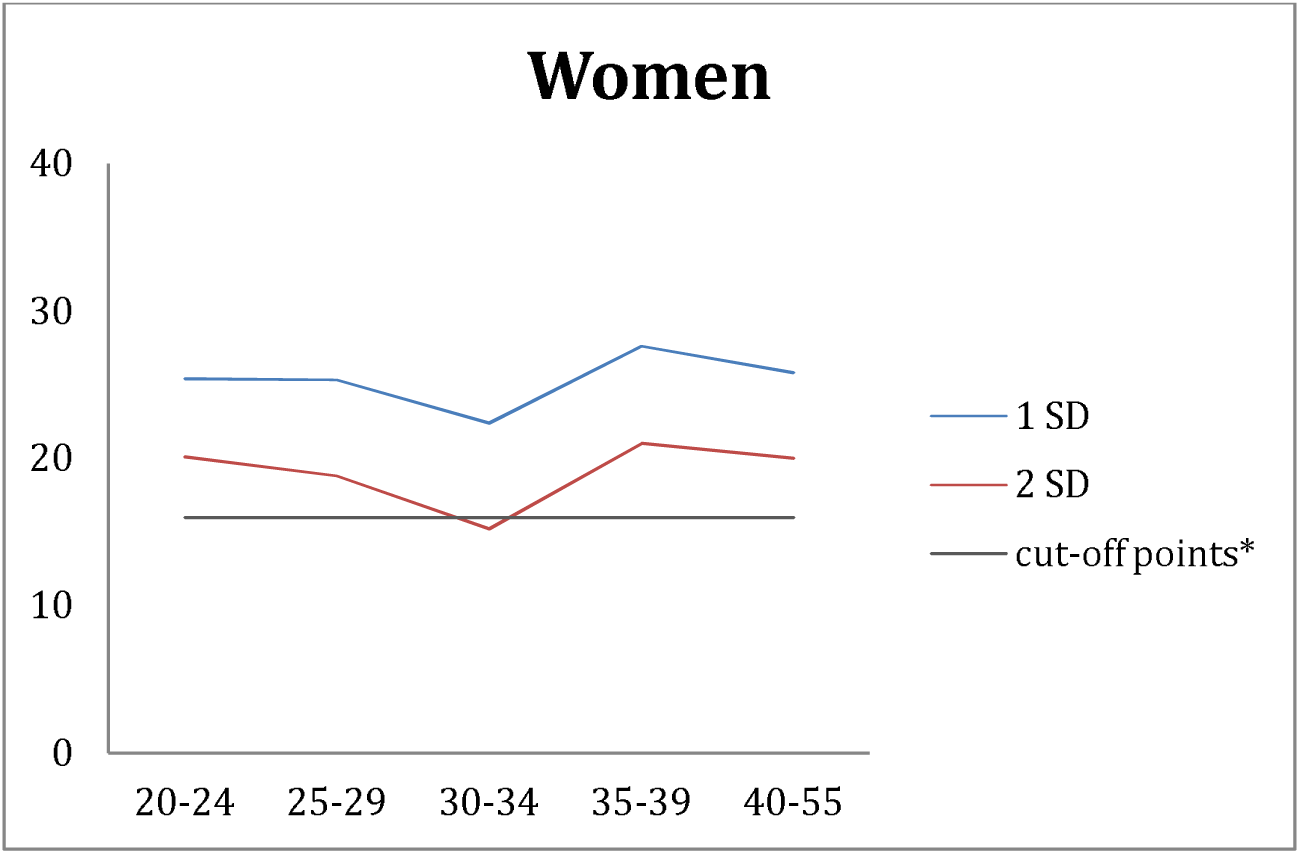
Distribution of women workers according to the mean grip strength values (below 1 SD and 2 SD), and the relation to the cut-off, according to the age groups.

The age group of 35-39 in the men’s group had the values of 1 SD and 2 SD, respectively, 42.7 kg and 34.6 kg. The lowest values of 1 SD and 2 SD found in the group of men are in the 45-57 age group with 38.8 kg and 29.9 kg, 1 and 2 SD respectively. However, no age groups in the men’s group are below the cut-off line (see fig 3b).

**Figure 3b.**
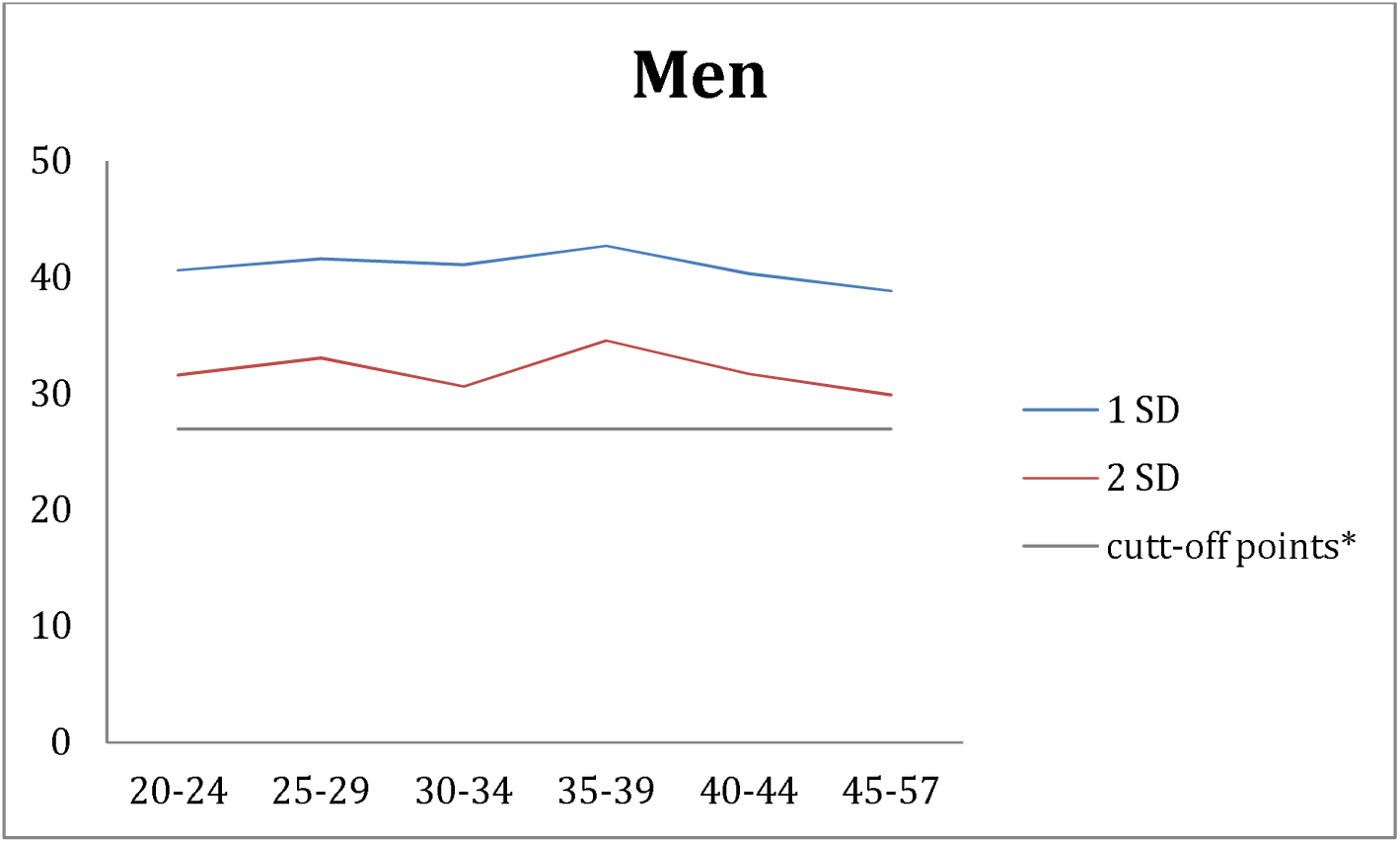
Distribution of men’s workers according to the mean grip strength values (below 1 SD and 2 SD), and the relation to the cut-off, according to the age groups.

The normative values for Portuguese workers are presented in table 1 and 2, separately for women and men. The women workers were divided into 5 age groups (20-24, 25-29, 30-34, 35-39, 40-55) and in the men workers were distributed by 6 age groups (20-24, 25-29, 30-34, 35-39, 40-44, 45-57). In the group of women, we chose to separate into five age groups, because the population over 40 years were not in large numbers.

**Table 1.**
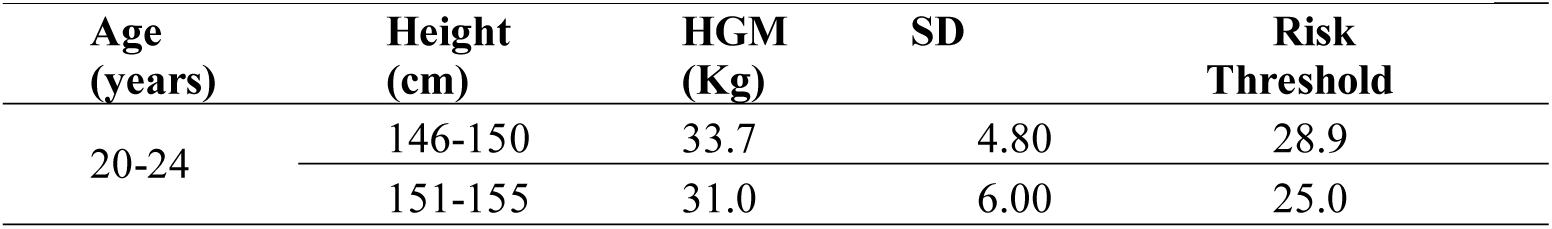
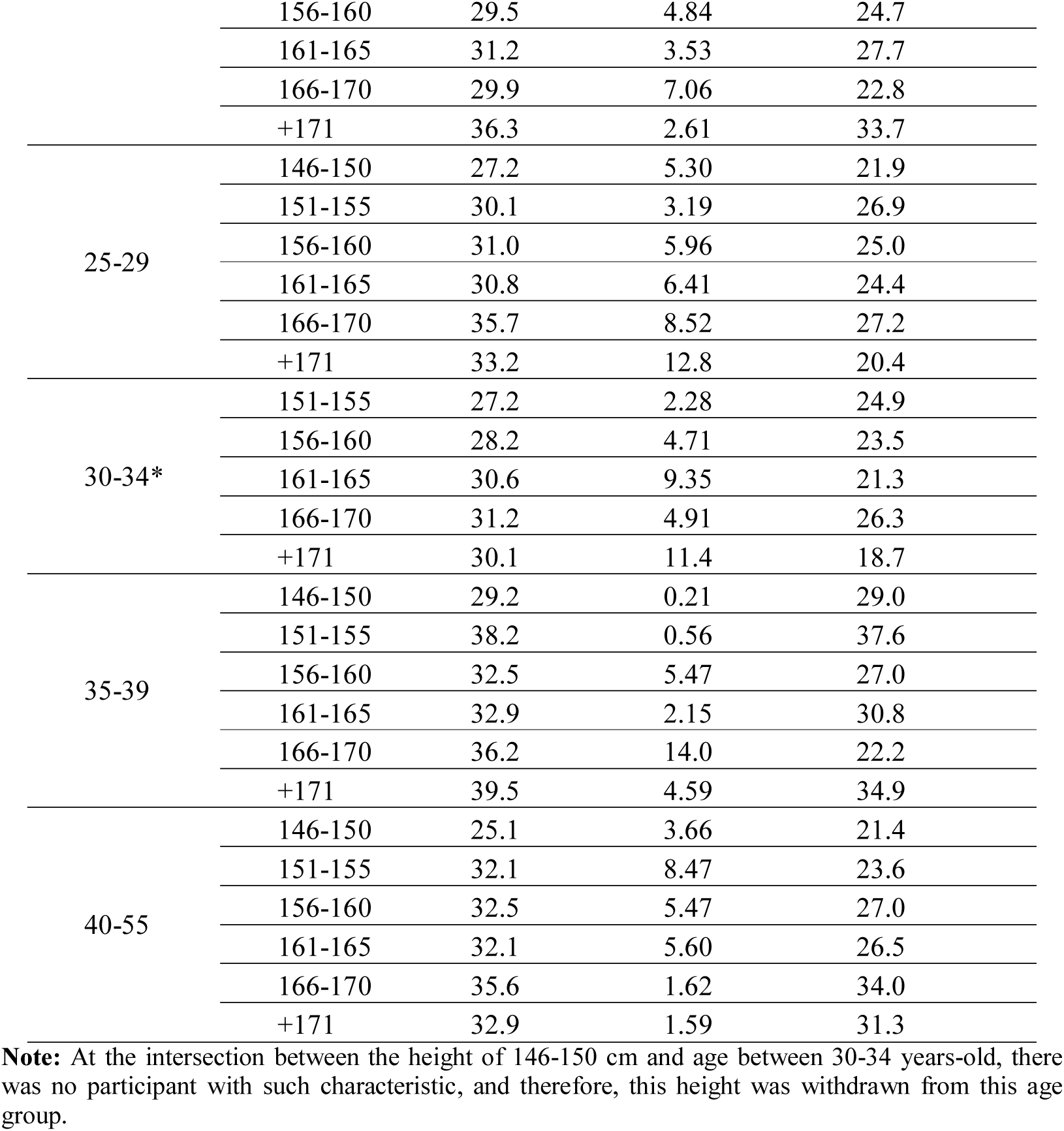
Normative reference values of Handgrip Strength for women

**Table 2.**
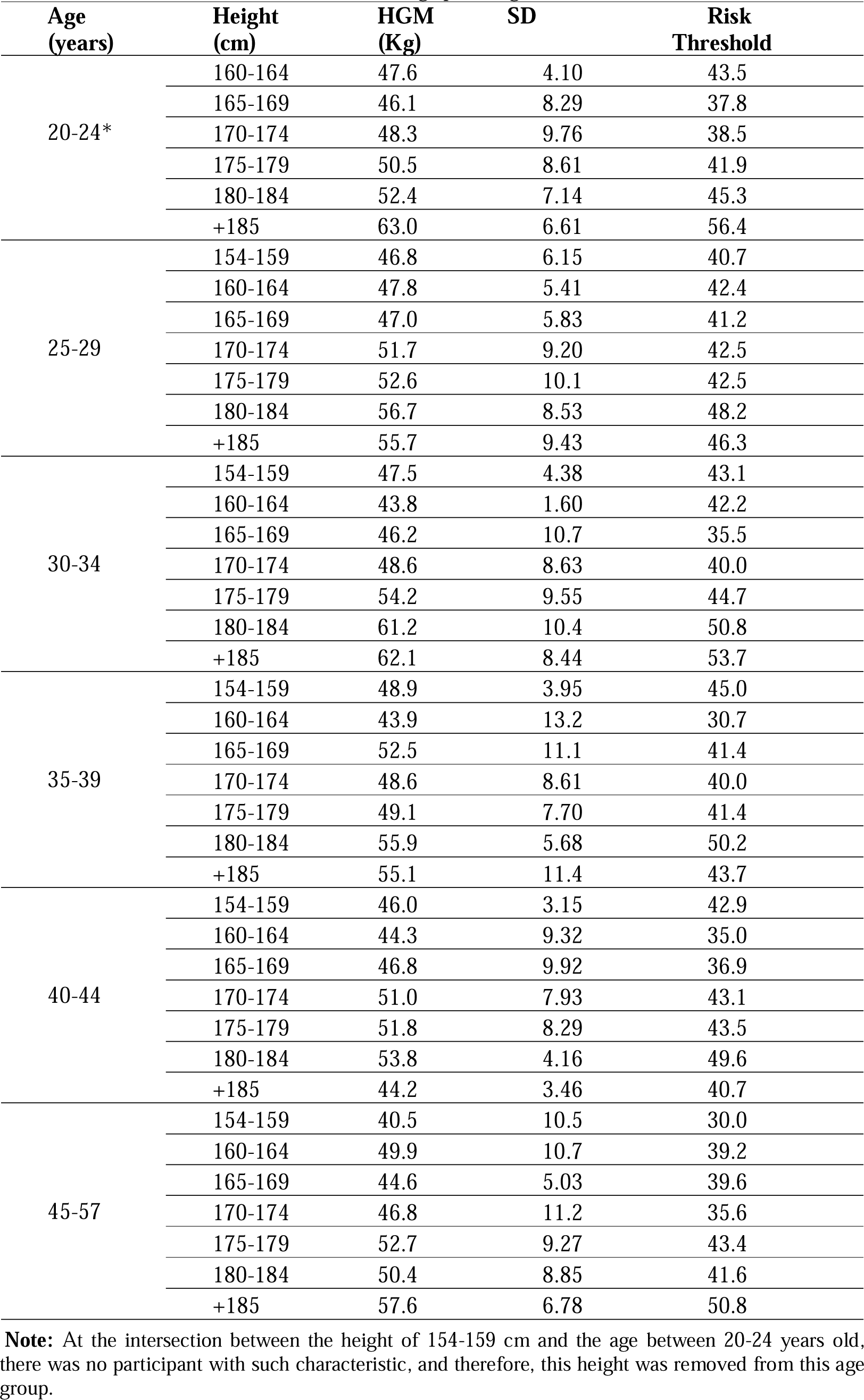
Normative reference values of Handgrip Strength for men

The maximum value of handgrip strength in the group of women is in the age group of 35-39 years at a height greater than (+) 171 cm, with 39.5 kg. Moreover, the minimum value appeared in the 40-55 age group at a height between 146-150cm, with 25.1kg (Table 1).

The maximum value of handgrip strength in the group of women is in the age group of 35-39 years at a height greater than (+) 171 cm, with 39.5 kg. Moreover, the minimum value appeared in the 40-55 age group at a height between 146-150cm, with 25.1kg (Table 1).

The maximum value of the handgrip strength was 63.0 kg for the men workers in the age group of 20-24 years and height higher than (+) 185 cm. The minimum value was about 40.5 kg in the age group of 45-57 years at the height 154-159cm (Table 2).

Table 3 presents the proportion of workers, according to age and height groups, of both genders, stratified by standard deviations, from the Z-standardized values of the handgrip strength, obtained through the Ordinary least square regression models.

**Table 3.** Distribution of workers by gender and age groups according the height –z-Standardized handgrip strength

|  | **n Male** | **%** | **n Female** | **%** |
| --- | --- | --- | --- | --- |
| Reference group sM(+/-0.5SD) | 182 | 40.9 | 67 | 42.7 |
| (1) 0.5SD <1.0 SD below sM | 68 | 15.3 | 28 | 17.8 |
| (2) 1.0 SD <1.5 SD below sM | 42 | 9.40 | 9 | 5.70 |
| (3) 1.5 SD <3.0 SD below sM | 21 | 4.70 | 10 | 6.40 |
| (4) 0.5 SD <1.0 SD above sM | 65 | 14.6 | 15 | 9.60 |
| (5) 1.0 SD <1.5 SD above sM | 40 | 9.00 | 17 | 10.8 |
| (6) 1.5 SD <3.0 SD above sM | 27 | 6.10 | 11 | 7.00 |
| Total | 445 | 91 | 157 | 100 |

**Note:** The sample consisted of men and women between the ages of 20 and 57 years. In the men's group, the body height has restricted in 160-200 cm. And the women group the body height is restricted from 146 to 181 cm. And the HGS measured is limited between 10 and 75 kg. The standardized handgrip strength was obtained from Ordinary least square and using the z-standardized residuals.

It was observed that the largest number of workers are in the reference group, and in the group of men only 4.7% of the sample of the workers are between 1.5 - <3.0 SD, below the mean values. For women workers only 6.4% are between 1.5 - <3.0 SD below the mean values for gender (Table 3).

## Discussion

### Main Findings

The study established normative values of handgrip strength of the Portuguese workers in the automotive industry in order to identify low grip strength limits, as a predictor of triggers disabilities such as frailty syndrome and sarcopenia.

The limit values were reported and stratified by sex, age, and body height. Stratification by height is essential as it has an influential role in handgrip strength. There are reports in studies that every 10 cm body height can lead to a 2-4 kg increase in handgrip strength (17).

In the automotive industry, or in other occupational settings, the use of height measurement can be an essential factor when designing work conditions tailored to aged workers. For example, in activities that require gripping of the objects above the head level, it would be appropriate to change the layout or work plan height in order to allow senior workers to perform them without constrains. Such preventive measure could prevent the emergence of musculoskeletal disorders and even the early onset of sarcopenia due to microtrauma developed by the impact of task demands (25–28).

Another interesting finding in the present study has found in the group of women between 30 and 34 years old, where there was a 4 kg drop in handgrip strength when compared to women in the age group of 35 to 39 years. This situation can be explained by the seniority in the company, where the age group that comprised the highest average peak force (35-39 years) was in the company for only two (2) years and was therefore less impacted by work demands, which remain the same for young and older workers (8,11,25,27,29).In the group of men, the decline is in the age group of 40-57 years, which was expected, since it is in this age group that the most characteristic decline occurs when compared to the other age groups (15,16,21,30,31). To our knowledge, our study is the first to produce normative grip strength values for the Portuguese workers in automotive industry, so we chose to compare the results of our study with 4 others previously published international studies. We considered the differences in mean peak values from previous studies with our average grip strength values, expressed as a percentage of our value and / or in kg.

In the first previously published study conducted in the German population, between the ages of 17 and 90 years, in both sexes, compared to the present study, the group of German men has a mean peak grip strength of 2% more than the male population of the present study. When compared to the present study, Portuguese women are 0.5% below the average peak force of the German women group (17).

The second study conducted with the English population, where 12 major reviews were compiled for the creation of the normative values of handgrip strength. The values of the peak handgrip strength were different when compared to the present study. Both the English men and women groups are 3% below the mean peak strength value observed in the Portuguese workers (21).

In a study conducted in the American population, normative values of handgrip strength were determined in the seven age groups. The handgrip strength peak of the American male population was 3.2% higher than the present men population. In the women’s group, when compared to the current study group, the average grip strength was 1.7% lower in all age groups (20).

By comparing the statistics from America, English, and German, with the population of the present study, we can infer that small differences in strength could be closely related to the type of activity (work demands) developed in the automotive sector. Effectively, there are several high demanding tasks, characterized by handling loads, unfavorable static postures, repetitive upper limb movements of force, influence on the occurrence of microtrauma surgery in the wrist, hand and elbow regions, interfering with the handgrip strength (25,29,32–34).

In another study of 187 male workers in the English automotive industry, the average peak handgrip strength was found in the age group 40-44 years with 47kg grip strength (35). Thus, the male population of the present study has 6 kg more average peak grip strength than the English car industry population. We can infer from the report of the author of the fourth study, that the situation of lower grip strength may be associated with the lack of calibration of the dynamometers used (35,36).

Our study has shown that grip strength has a peak increase in early adulthood (20-24 years) in both genders, and then continues to enter a not so pronounced decline with advancing age (from the age of 40). However, as explained above, the group of women (30-34 years) due to occupational exposure had a more pronounced decline (27).

Also this study found a high prevalence of weak grip strength based on 1 and 2 SD and cut-off points based on (37). In the 30-34 age group for women and in the 45-57 age group with 15.2 kg for women and 29.9 kg for men, thus producing more discriminatory cutoff values for grip strength by 40% for women (30-34 years) and 23.2% in the group of men aged 45-57 years (see fig 3a and 3b). The cut-off could be a significant ally for the automotive industry, where these age groups in both genders should be monitored more frequently to prevent the onset of sarcopenia and frailty syndrome (4).

## Conclusion

Normative values can help delineate the career path of workers because they portray risk values according to age, height, and gender. And they can also help in adjusting the morphological and strength characteristics of the worker with the task to be performed, as an example of work above head level. Also, the collaborators of the present study, in all the age groups, of the group of women as well as of the men, did not present sarcopenia effects neither the frailty syndrome.

## Data Availability

Relevant data is exposed in the article and complementary data will be available for support.

https://doi.org/10.5061/dryad.7m0cfxpq8

